# Age-dependent associations of sleep duration and insomnia symptoms with cognitive decline across five decades of adult life: a multi-cohort longitudinal study

**DOI:** 10.64898/2026.09.25.26363056

**Authors:** Yi Fang, Allysa Quick, Nina Oryshkewych, Nicole Fernandez, Andrew S Lim, Lisa L Barnes, David A Bennett, Andrew Steptoe, Rebecca Hardy, Séverine Sabia, Meredith L Wallace, Yue Leng

## Abstract

**Background:** Associations of sleep duration and insomnia symptoms with cognitive decline are inconsistent, and whether they represent modifiable risk factors or early manifestations of neurodegeneration may depend on life stage. We examined how sleep-cognition associations vary with age (44 to >90 years).

**Methods:** We studied 16,884 participants from three cohorts with harmonised measures. In the English Longitudinal Study of Ageing (ELSA: n=9,473, median age 64, range 50 to ≥90), linear mixed-effects models examined age-specific differences in cognitive change (recall and fluency) associated with sleep duration, difficulty falling asleep, nocturnal awakenings, incorporating sleep-by-age-by-time interactions, with initial adjustment for sociodemographic factors and further adjustment for health and lifestyle factors. Sleep characteristics showing consistent age-dependent associations in ELSA were further evaluated in Whitehall II (n=5,580, median age 55, range 44 to 68) and the Rush Memory and Aging Project (MAP: n=1,831, median age 80, range 53 to 102).

**Findings:** In ELSA, difficulty falling asleep ≥1 time/week (vs <1 time/week) was associated with less favourable 5-year change in recall at age 50 (difference −0·12 SD, 95% CI −0·20 to −0·04), with the association attenuating and reversing towards more favourable change at older ages (p-interaction=0·009 [linear], 0·016 [quadratic]). These persisted after further adjustment for health and lifestyle factors. Short sleep (≤6 h) showed similar patterns that attenuated after further adjustment. Similar age gradients were observed for difficulty falling asleep in Whitehall II and MAP. There was no evidence of age modification on long sleep and nocturnal awakenings.

**Interpretation:** The cognitive implications of sleep disturbances vary by life stage. In midlife, difficulty falling asleep was associated with faster cognitive decline, supporting its role as a modifiable risk factor. In advanced age, the reversal is consistent with diminished arousal capacity accompanying prodromal dementia.

**Funding:** US National Institute on Aging

**Research in context:** *Evidence before this study:* We searched PubMed and Web of Science from database inception to Jan 1, 2026, using terms related to sleep, sleep duration, insomnia, cognitive decline, dementia, and ageing, without language restrictions. Findings on sleep and cognitive ageing are inconsistent and vary by population studied. In the few prospective studies of midlife populations, short sleep and insomnia symptoms have been linked to increased dementia risk, but evidence remains limited. In older populations, findings are more mixed, with several studies reporting null associations and others reporting counterintuitive associations between greater sleep complaints and better cognitive outcomes. Few studies span a sufficient age range to formally test age as an effect modifier; those that did were often limited by modest sample sizes, binary age stratification, or failure to model time within the interaction.

*Added value of this study:* To our knowledge, this is the first multi-cohort study to examine age as a modifier of the association between sleep and longitudinal cognitive trajectories across general population spanning midlife to the oldest old age. In a nationally representative UK cohort, we show that difficulty falling asleep has robust age-dependent associations with cognitive decline: faster decline in midlife, attenuation through later adulthood, and reversal in the oldest old. Short sleep duration shows a similar but less robust age-dependent pattern, partly explained by health-related and lifestyle factors. These findings were replicated across two independent cohorts in the UK and US with complementary age ranges.

*Implications of all the available evidence:* The available evidence supports age-specific interpretation of sleep disturbances in relation to cognitive health. In midlife, difficulty falling asleep is associated with faster cognitive decline, supporting its potential as a modifiable target for intervention. In the oldest old, difficulty falling asleep may reflect lower sleep propensity from preserved wake-promoting function, consistent with converging evidence linking increased sleepiness to neurodegeneration.

## Introduction

Sleep and cognitive function both change substantially across the adult life course,^1,2^ and growing evidence points to a bidirectional relationship between the two: sleep disturbances may contribute to neurodegenerative processes, but may also arise as early manifestations of underlying neuropathology.^3^ Dementia develops over a prolonged preclinical phase^4^ during which pathological changes—including in brain regions regulating arousal and sleep–wake stability^5^—accumulate decades before cognitive symptoms emerge, such that alterations in sleep duration and quality may arise during this preclinical period.^6^ This bidirectional relationship has fuelled debate over whether sleep disturbances act primarily as modifiable risk factors or prodromal markers of dementia, a distinction that may vary by stage of life.^7^

A life-course perspective spanning midlife and later life provides a framework to clarify this debate. In midlife, before substantial neurodegenerative pathology has accumulated,^8^ sleep disturbances may primarily reflect upstream risk processes contributing to subsequent cognitive decline. In later life, ongoing neurodegenerative pathology may progressively impair wake-promoting systems,^9^ increasing sleep propensity and potentially leading to fewer reported insomnia symptoms, such that their absence may no longer indicate good sleep health but rather reflect underlying neuropathology. Consistent with this framework, studies in midlife populations generally link short sleep duration and insomnia symptoms to worse cognition and higher dementia risk,^10–12^ whereas findings in older adults have been less consistent, including null and inverse associations.^13–18^ However, few studies span both midlife and later life with adequate sample sizes and sufficient follow-up, and fewer still formally test whether age modifies the longitudinal association between sleep and cognitive decline.^19–21^ Understanding how the cognitive implications of sleep disturbances vary across the adult life course could help clarify when sleep may represent potentially modifiable risk factors and when they may instead serve as markers of underlying neurodegenerative change.

We therefore investigated age-dependent associations of baseline sleep duration and insomnia symptoms with subsequent cognitive trajectories in more than 16,000 adults from three longitudinal cohorts in the UK and USA. We selected the English Longitudinal Study of Ageing (ELSA) for the primary analysis because its large sample and broad age distribution at sleep assessment, spanning ages 50 to ≥90 years, allowed us to examine age modification within a single cohort. Using harmonised measures and analytic methods, we then assessed whether the age-related gradient in the association between difficulty falling asleep and cognitive decline observed in ELSA was also evident in Whitehall II (WHII), where baseline sleep was assessed predominantly in midlife, and the Rush Memory and Aging Project (MAP), comprising predominantly older adults. We hypothesised that short sleep duration and more frequent insomnia symptoms would be more strongly associated with faster cognitive decline in midlife.

## Methods

### Data sources and study participants

We analysed data from three longitudinal cohort studies: the ELSA (UK), WHII (UK), and the MAP (USA). ELSA is a population-based study of adults aged ≥50 years living in England, initiated in 2002. We defined wave 4 (2008–2009) as baseline, as it was the first wave to include detailed self-reported sleep measures alongside standardised cognitive testing. Cognition was assessed at waves 5 (2010–2011), 6 (2012–2013), 7 (2014–2015), 8 (2016–2017), and 9 (2018–2019).

WHII is an occupational cohort of British civil servants aged 35–55 years at recruitment (1985–1988). Wave 5 (1997–1999), when sleep and cognitive measures were first jointly administered, served as baseline. Follow-up cognitive assessments were conducted at waves 7 (2002–2004), 9 (2007–2009), 11 (2012–2013), and 12 (2015–2016).

MAP is an ongoing community-based cohort initiated in 1997, with rolling enrolment of older adults without known dementia from retirement communities, senior housing facilities, and individual homes in the greater Chicago area. Participants undergo annual cognitive assessments. Baseline was defined as the first visit at which both sleep and cognition were examined.

Across cohorts, participants were included if they had at least one baseline sleep measure, a cognitive assessment, and data on key demographic covariates. Individuals with prevalent dementia at baseline were excluded. Cohort-specific dementia ascertainment procedures are described in the **Supplement**. All studies received ethical approval from relevant institutional review boards, and participants provided written informed consent.

### Sleep duration and insomnia symptoms

In ELSA, we examined habitual sleep duration, difficulty falling asleep, and nocturnal awakenings. Sleep duration was categorised as short (≤6 hours), reference (>6 to <8 hours), or long (≥8 hours). Difficulty falling asleep and nocturnal awakenings were assessed by frequency over the past month and dichotomised as low (<1 time/week) or high (≥1 time/week).

External-cohort analyses in Whitehall II and MAP focused on difficulty falling asleep because it showed the most consistent evidence of age-dependent associations across cognitive outcomes and adjustment models in ELSA. Responses were harmonised to distinguish less frequent from more frequent symptoms across cohorts (**Table S1**). In Whitehall II, high frequency was defined as symptoms reported on ≥4 days during the past month, approximately equivalent to ≥1 time/week. In MAP, high frequency comprised responses of “sometimes,” “often,” or “very often,” and low frequency comprised “never” or “rarely.”

### Cognitive measurement

In ELSA, cognitive outcomes were word recall and verbal fluency. Corresponding measures were examined in Whitehall II and MAP, alongside additional cohort-specific cognitive domains in secondary analyses (**Table S2**).

Word recall was assessed using immediate recall of a 10-word list in ELSA, a 20-word list in Whitehall II, and a CERAD word-list recall task in MAP. Verbal fluency was assessed using animal naming in ELSA and MAP and a composite of category (animals) and letter (S) fluency in Whitehall II. Verbal fluency was not assessed at ELSA wave 6. Within each cohort, test scores were standardised using the baseline sample mean and standard deviation, with the same reference values applied to subsequent assessments. Higher scores indicated better performance.

### Covariates

Cohort-specific definitions and harmonised covariates are provided in **Table S3**. Covariates were selected a priori based on their potential associations with sleep and cognition. Sociodemographic covariates included baseline age, sex, race and ethnicity (self-reported; categorised according to cohort-specific classifications: White and other racial or ethnic groups in ELSA and WHII; non-Hispanic White and other racial or ethnic groups in MAP), education (O-level or below vs A-levels or above, or equivalent), marital status (married or cohabitating vs other), and employment status (employed, retired, unemployed; not collected in MAP, where most participants were retired). Health-related covariates included hypertension, diabetes, cardiovascular disease, depressive symptoms, and sleep medication use (available in WHII and MAP but not in ELSA). Lifestyle covariates included smoking status (never vs ever), alcohol use (any vs none in the past year), physical activity (cohort-specific categorisation), and body mass index (<20, 20–24·9, 25–29·9, ≥30 kg/m^2^).

### Statistical Analyses

Participant characteristics were summarised using median (interquartile range [IQR]) for continuous variables and n (%) for categorical variables.

Linear mixed-effects models were used to estimate longitudinal trajectories of cognitive z scores, with participant-specific random intercepts and slopes for follow-up time. Follow-up time was modelled using linear and quadratic terms to capture non-linear trajectories, supported by model fit statistics (Bayesian Information Criterion and likelihood ratio tests). To assess effect modification by baseline age, models included a three-way interaction between sleep, baseline age, and follow-up time, along with all corresponding lower-order terms.

Three progressively adjusted models were specified. Model 1 adjusted for sociodemographic covariates. Model 2 additionally adjusted for health-related and lifestyle factors. Model 3 further adjusted for *APOE* e4 carrier status where available (WHII and MAP). Models 2 and 3 were fitted in complete-case samples for each model; consistency of estimates across models was examined.

To quantify age-dependent associations, marginal mean cognitive z scores were estimated at prespecified baseline ages, and differences in cognitive change across sleep categories were then derived over cohort-specific follow-up intervals. The selected baseline ages reflected cohort-specific age distributions: ELSA (50, 55, 60, 65, 70, 75, 80, 85, and 90 years), WHII (45, 50, 55, 60, and 65 years), and MAP (70, 75, 80, 85, and 90 years). Follow-up intervals were tailored to each cohort’s available follow-up across ages: 5-year and 10-year change in ELSA, 15-year change in WHII, and 5-year change in MAP. Because intervals differ and trajectories include non-linear components, cross-cohort comparisons focus on the direction and age gradient of associations rather than absolute effect sizes. Model-predicted cognitive trajectories across baseline ages and sleep categories were visualised to illustrate age-dependent patterns.

Because MAP participants were enrolled on a rolling basis, follow-up was divided into consecutive 5-year waves to maximise use of longitudinal data. The start of each wave was treated as a new baseline, and participants could contribute up to four non-overlapping waves, depending on follow-up duration.

We conducted sensitivity analyses to assess the robustness of the main findings. In ELSA, we examined alternative categorisations of sleep duration and insomnia symptoms after excluding participants who developed dementia during follow-up (n=420), to assess sensitivity to exposure cut-points and minimise potential bias from incipient dementia affecting self-reported sleep. To account for potential informative dropout, we additionally fitted Bayesian joint models of cognitive trajectories and time to loss to follow-up across all three cohorts, focusing on difficulty falling asleep in relation to recall and verbal fluency. The longitudinal submodel retained the fixed- and random-effects structure of the corresponding Model 1 mixed-effects analysis, and both submodels included the cohort-specific Model 1 covariates. We compared the direction and magnitude of the sleep-by-age-by-time interaction estimates between the mixed-effects and joint models and assessed whether interactions identified in the mixed-effects analyses remained supported by posterior estimates with 95% credible intervals.

All analyses were conducted in R (version 4·4.3) using the lme4, lmerTest, emmeans, and JMbayes2 packages. Two-sided p values <0·05 were considered statistically significant. Formal correction for multiple comparisons was not applied; instead, given the limited power of higher-order interactions,^22^ we prioritised replication of age-dependent patterns across cohorts and cognitive outcomes over p-value thresholds. Interpretation focused on the direction, magnitude, and consistency of effect estimates.

### Role of the funding source

The funder of the study had no role in study design, data collection, data analysis, data interpretation, or writing of the report.

## Results

### Sample characteristics

Participant flow is shown in **Figure S1**. In ELSA, 9,473 participants were included in the primary analysis, with 7,506 retained for Model 2 after accounting for additional covariates. Corresponding numbers were 5,580 and 4,155 in WHII. In MAP, 1,831 participants contributed to the primary analysis and 1,812 to Model 2. For Model 3, analyses were conducted among participants with available *APOE* e4 data, yielding 3,239 in WHII and 1,479 in MAP. Because MAP follow-up was structured into consecutive non-overlapping 5-year observational windows, 783, 251, and 72 participants contributed data to a second, third, and fourth window, resulting in more analytic observations than unique participants.

Baseline characteristics are presented in **Table 1**. Age distributions varied across cohorts (**Figure S2**), with median (IQR; range) baseline age of 64 (58–73; 50–≥90) years in ELSA, 55 (50–61; 44–68) years in WHII, and 80 (75–85; 53–102) years in MAP. After accounting for 5-year windows, the median (IQR) age at the start of MAP participant-wave observations was 82 (76–87) years. Follow-up duration also varied across cohorts and declined with advancing baseline age within each cohort (**Table S4**). Median (IQR) follow-up time was 9·7 (4·0–10·1) years in ELSA, 17·6 (15·4–18·1) years in WHII, and 4·7 (1·3–8·3) years in MAP. Baseline characteristics by analytic samples and MAP observation windows are presented in **Table S5-S7**. Participants included in Models 2 and 3 differed from those excluded due to missing data, indicating differences in analytic samples across models.

**Table 1.** Baseline characteristics of participants in the ELSA, Whitehall II, and MAP cohorts.

|  | **ELSA (n=9473)** | **Whitehall II (n=5580)** | **MAP (n=1831)** |
| --- | --- | --- | --- |
| Age at baseline, years | 64 (58–73) | 55 (50–61) | 80 (75–85) |
| Year of birth | 1943 (1935–1950) | 1943 (1937–1947) | Not available |
| Length of cognitive follow-up, years | 9·7 (4·0–10·1) | 17·6 (15·4–18·1) | 4·7 (1·3–8·3) |
| Number of cognitive follow-up visits | 5 (3–6) | 5 (4–5) | 5 (2–9) |
| Sex | .. | .. | .. |
| Male | 4219 (44·5%) | 4031 (72·2%) | 460 (25·1%) |
| Female | 5254 (55·5%) | 1549 (27·8%) | 1371 (74·9%) |
| Race and ethnicity | .. | .. | .. |
| White in UK cohorts;<br>non-Hispanic White in MAP | 9205 (97·2%) | 5132 (92·0%) | 1608 (87·8%) |
| Other racial or ethnic groups | 268 (2·8%) | 448 (8·0%) | 223 (12·2%) |
| Education ≥A-level or equivalent | 3671 (38·8%) | 3582 (64·2%) | 1335 (72·9%) |
| Married or partnered | 6522 (68·8%) | 4264 (76·4%) | 720 (39·3%) |
| Employment status | .. | .. | Not available |
| Employed | 3175 (33·5%) | 3735 (66·9%) | Not available |
| Retired | 5165 (54·5%) | 1484 (26·6%) | Not available |
| Unemployed | 1133 (12·0%) | 361 (6·5%) | Not available |
| Hypertension | 4937 (52·4%) | 1923 (34·7%) | 1235 (68·1%) |
| Diabetes mellitus | 925 (9·8%) | 345 (7·0%) | 248 (13·5%) |
| Cardiovascular disease | 853 (9·1%) | 776 (13·9%) | 279 (15·2%) |
| Depressive symptoms | 2540 (27·0%) | 680 (12·4%) | 182 (9·9%) |
| Ever smoker | 5709 (60·7%) | 2775 (50·0%) | 785 (42·9%) |
| Alcohol use in the past year | 7239 (78·2%) | 5274 (95·9%) | 898 (49·0%) |
| Body mass index, kg/m<sup>2</sup> | 27·4 (24·7–30·9) | 25·6 (23·5–28·0) | 26·6 (23·9–30·2) |
| Sleep medication use | Not available | 191 (3·4%) | 503 (27·5%) |
| APOE ε4 carrier | Not available | 1202 (28·1%) | 344 (23·1%) |
| Sleep duration | .. | .. | .. |
| 6–8 h | 3441 (36·4%) | 2358 (42·5%) | 595 (32·5%) |
| ≤6 h | 3153 (33·4%) | 2291 (41·3%) | 500 (27·3%) |
| ≥8 h | 2852 (30·2%) | 896 (16·2%) | 736 (40·2%) |
| Difficulty falling asleep | 2621 (27·7%) | 981 (17·8%) | 793 (43·3%) |
| Nocturnal awakenings | 6282 (66·3%) | 2508 (45·6%) | 1186 (64·8%) |

Data are presented as median (IQR) or n (%). Race and ethnicity were self-reported and categorised according to cohort-specific classifications (White in ELSA and Whitehall II; non-Hispanic White in MAP). Difficulty falling asleep and nocturnal awakenings are reported as the proportion of participants with symptoms occurring ≥1 time/week in ELSA and Whitehall II and sometimes/often/very often in MAP. APOE ε4 carrier status and sleep medication use were available in Whitehall II and MAP. Employment status was available in ELSA and Whitehall II. ELSA=English Longitudinal Study of Ageing; MAP=Rush Memory and Aging Project.

### ELSA

Three-way interactions between sleep, baseline age, and follow-up time are shown in **Table 2**. Difficulty falling asleep showed significant interactions with baseline age and follow-up time for word recall in both Models 1 and 2, across both linear and quadratic time terms. For verbal fluency, interaction was primarily observed for the linear term. Model-estimated differences in cognitive decline from Model 1 are summarised in **Figure 1**. At age 50 years, reporting difficulty falling asleep ≥1 time/week was associated with a less favourable difference in 5-year recall change compared with those reporting symptoms less than once per week (between-group difference −0·12 SD, 95% CI −0·20 to −0·04). This effect attenuated with increasing age, approaching null by around age 70 years, and reversed at older ages, consistent with less cognitive decline among those reporting frequent symptoms. Similar age-dependent patterns were observed for verbal fluency: differences were modest in midlife but became increasingly positive after approximately age 70 years, consistent with less decline in verbal fluency among those reporting symptoms ≥1 time/week compared with those reporting less frequent symptoms. Ten-year estimates showed a similar but attenuated gradient. These patterns remained after further adjustment in Model 2 (**Figure S3**). Predicted trajectories from Model 2 (**Figure 2**) showed lower cognitive scores among those reporting frequent difficulty falling asleep at younger baseline ages, little difference around 65–75 years, and higher scores at older ages (≥80 years).

**Figure 1.**
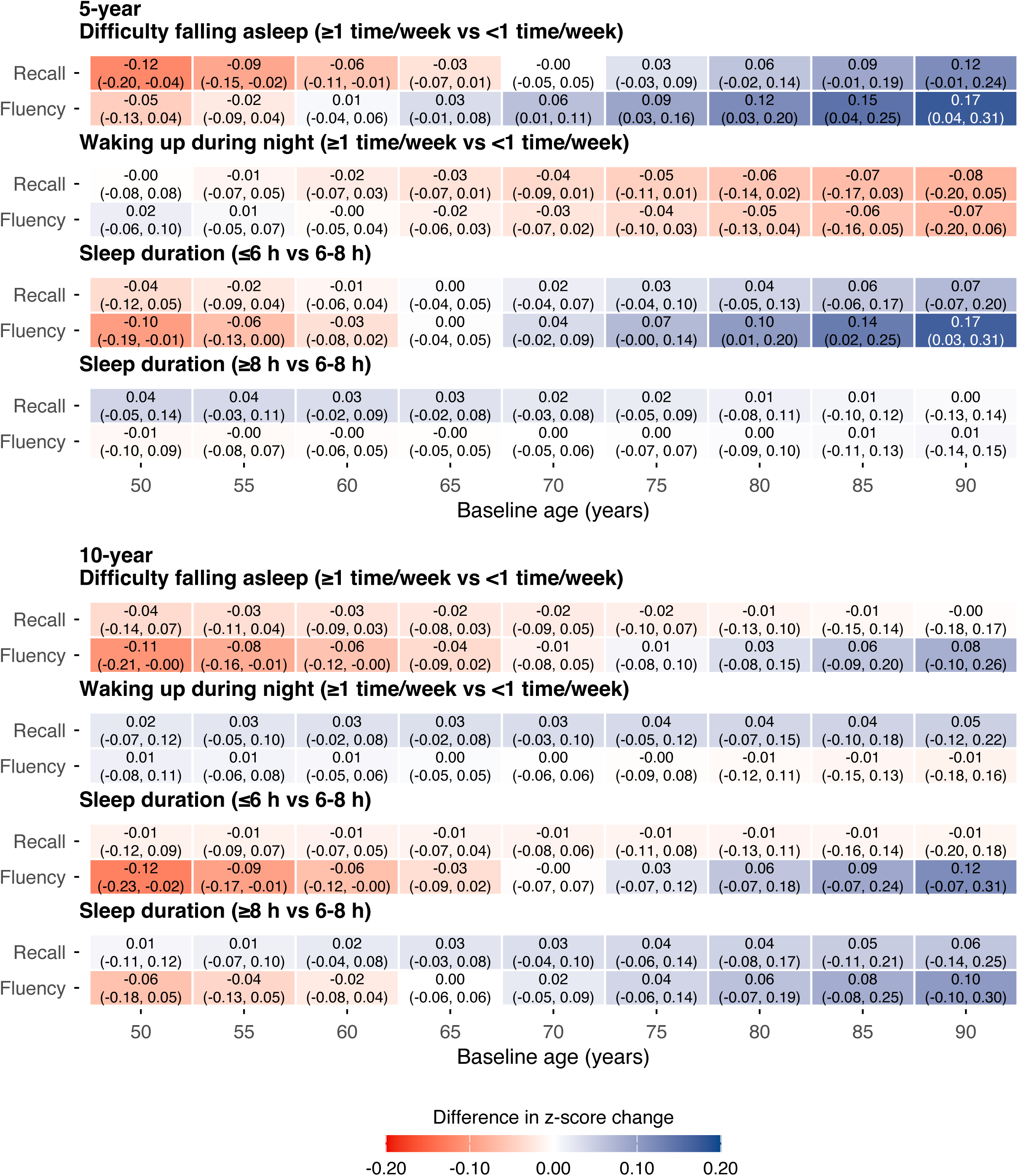
Associations of sleep measures with change in verbal fluency and word recall in ELSA, Model 1. Heatmaps show estimated differences (95% CI) in 5-year and 10-year changes in cognitive z scores between sleep categories (difficulty falling asleep and nocturnal awakenings ≥1 time/week vs <1 time/week; sleep duration ≤6 h or ≥8 h vs 6–8 h) across baseline ages 50–90 years. Estimates were derived from marginal means predicted by linear mixed-effects models including three-way interactions between baseline age, sleep, and follow-up time, along with all lower-order terms, and adjusted for sex, race and ethnicity, education, marital status, and employment status. Random intercepts and slopes were included to account for within-individual correlations. Colours indicate the magnitude and direction of associations. Positive values (blue) indicate relatively less cognitive decline compared with the reference sleep category, whereas negative values (red) indicate relatively greater decline.

**Figure 2.**
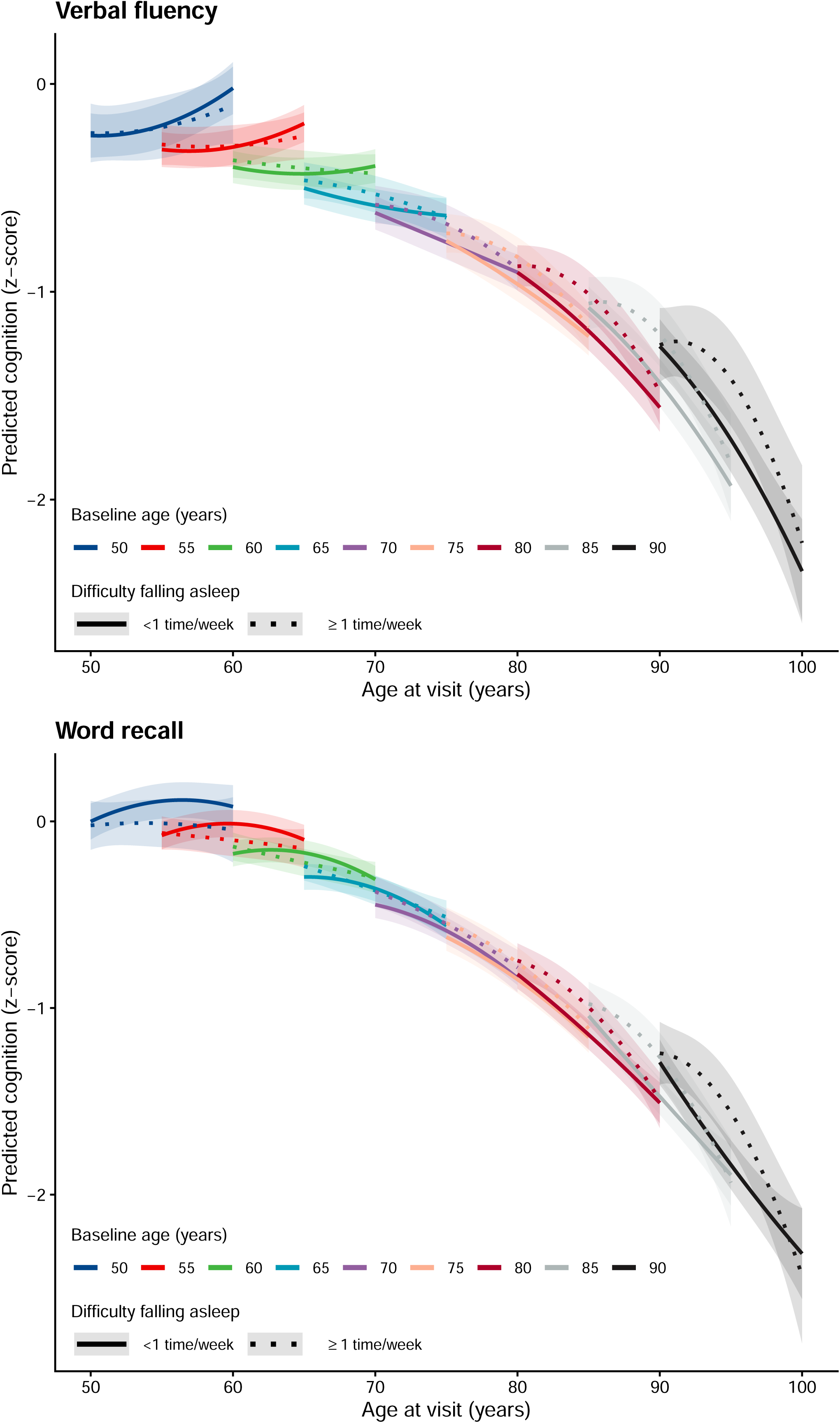
Predicted trajectories of verbal fluency and word recall by baseline age and difficulty falling asleep in ELSA, Model 2. Predicted cognitive trajectories (z scores) for verbal fluency and word recall by baseline age (50–90 years) and difficulty falling asleep (≥1 time/week vs <1 time/week) in the English Longitudinal Study of Ageing (ELSA). Lines represent estimated marginal means from mixed-effects models (Model 2); shaded areas indicate 95% confidence intervals. Colour indicates baseline age. Solid line indicates <1 time/week, dotted line indicates ≥1 time/week. Follow-up is truncated at 10 years. The model was adjusted for age, sex, race and ethnicity, education, marital status, employment status, body mass index, hypertension, diabetes, cardiovascular disease, depressive symptoms, smoking, alcohol use, and physical activity.

**Table 2.** Three-way interaction between baseline sleep, baseline age, and follow-up time on cognitive outcomes in ELSA.

|  |  |  | Model 1 |  | Model 2 |  |
| --- | --- | --- | --- | --- | --- | --- |
| Sleep variable | Cognitive outcome | Time component | Coefficient (95% CI) | p value | Coefficient (95% CI) | p value |
| Difficulty falling asleep | Fluency | Linear | 0.177 (-0.005 to 0.358) | 0.056 | 0.218 (0.015 to 0.420) | 0.035 |
| Difficulty falling asleep | Fluency | Quadratic | -0.130 (-0.319 to 0.059) | 0.18 | -0.171 (-0.381 to 0.039) | 0.11 |
| Difficulty falling asleep | Recall | Linear | 0.228 (0.056 to 0.400) | 0.009 | 0.275 (0.084 to 0.467) | 0.005 |
| Difficulty falling asleep | Recall | Quadratic | -0.219 (-0.398 to -0.041) | 0.016 | -0.265 (-0.462 to -0.068) | 0.008 |
| Sleep duration ($\leq 6$ h) | Fluency | Linear | 0.206 (0.013 to 0.400) | 0.036 | 0.100 (-0.114 to 0.314) | 0.36 |
| Sleep duration ($\leq 6$ h) | Fluency | Quadratic | -0.146 (-0.346 to 0.054) | 0.15 | -0.071 (-0.291 to 0.149) | 0.53 |
| Sleep duration ($\leq 6$ h) | Recall | Linear | 0.103 (-0.081 to 0.287) | 0.27 | -0.024 (-0.227 to 0.178) | 0.81 |
| Sleep duration ($\leq 6$ h) | Recall | Quadratic | -0.102 (-0.291 to 0.087) | 0.29 | 0.028 (-0.179 to 0.235) | 0.79 |
| Sleep duration ($\geq 8$ h) | Fluency | Linear | -0.027 (-0.229 to 0.176) | 0.80 | -0.117 (-0.342 to 0.108) | 0.31 |
| Sleep duration ($\geq 8$ h) | Fluency | Quadratic | 0.068 (-0.143 to 0.279) | 0.53 | 0.153 (-0.079 to 0.385) | 0.20 |
| Sleep duration ($\geq 8$ h) | Recall | Linear | -0.050 (-0.242 to 0.142) | 0.61 | -0.050 (-0.263 to 0.162) | 0.64 |
| Sleep duration ($\geq 8$ h) | Recall | Quadratic | 0.062 (-0.137 to 0.261) | 0.54 | 0.083 (-0.135 to 0.301) | 0.46 |
| Nocturnal awakenings | Fluency | Linear | -0.081 (-0.254 to 0.092) | 0.36 | -0.058 (-0.251 to 0.136) | 0.56 |
| Nocturnal awakenings | Fluency | Quadratic | 0.074 (-0.105 to 0.254) | 0.42 | 0.040 (-0.159 to 0.240) | 0.69 |
| Nocturnal awakenings | Recall | Linear | -0.080 (-0.244 to 0.084) | 0.34 | 0.015 (-0.168 to 0.198) | 0.88 |
| Nocturnal awakenings | Recall | Quadratic | 0.087 (-0.083 to 0.256) | 0.32 | -0.005 (-0.192 to 0.182) | 0.96 |

Coefficients represent three-way interaction terms between baseline sleep, baseline age, and follow-up time from linear mixed-effects models. Baseline age was centred at the cohort mean, and both baseline age and follow-up time were modelled per 10-year increase. Linear and quadratic terms correspond to interactions with the linear and squared follow-up time components, respectively.
Model 1 was adjusted for age, sex, race and ethnicity, education, marital status, and employment status. Model 2 was additionally adjusted for hypertension, diabetes, cardiovascular disease, depressive symptoms, smoking status, alcohol use, physical activity, and body mass index.
The reference category for sleep duration was 6–8 h. For sleep disturbance variables, estimates compare high-frequency symptoms ( $\geq 1$ time/week) with low-frequency symptoms ( $< 1$ time/week). Estimates are presented with 95% confidence intervals, and all p values are two-sided.

For sleep duration, short sleep (≤6 h vs 6–8 h) showed a significant age-dependent association with verbal fluency in Model 1 (**Table 2**). At age 50 years, short sleep was associated with a 0·10 SD greater 5-year decline in verbal fluency (95% CI −0·19 to −0·01), with attenuation to null by approximately age 65 years. Beyond this age, the direction of effect reversed. At age 80 years, short sleep was associated with 0·10 SD less decline (0·01 to 0·20), increasing to 0·14 SD (0·02 to 0·25) at age 85 and 0·17 SD (0·03 to 0·31) at age 90 (**Figure 1**). Ten-year estimates showed a similar pattern, although estimates at older ages were less precise. Adjustment for health behaviours and comorbidities in Model 2 attenuated evidence for age modification (**Figure S3**). No clear age modification was observed for long sleep duration (≥8 h) or nocturnal awakenings, with estimates remaining close to null across the age spectrum.

### Replication in WHII and MAP

In Whitehall II, where sleep was assessed predominantly in midlife, more frequent difficulty falling asleep was associated with greater predicted 15-year decline in fluency at younger baseline ages, with differences attenuating at older ages (**Figure 3**; **Figures S4–S5**). In Model 3, the difference was −0·13 SD (95% CI −0·24 to −0·01) at age 45. Despite this gradient, the quadratic three-way interaction was significant only in Model 1 (p=0·021; **Table S9**).

**Figure 3.**
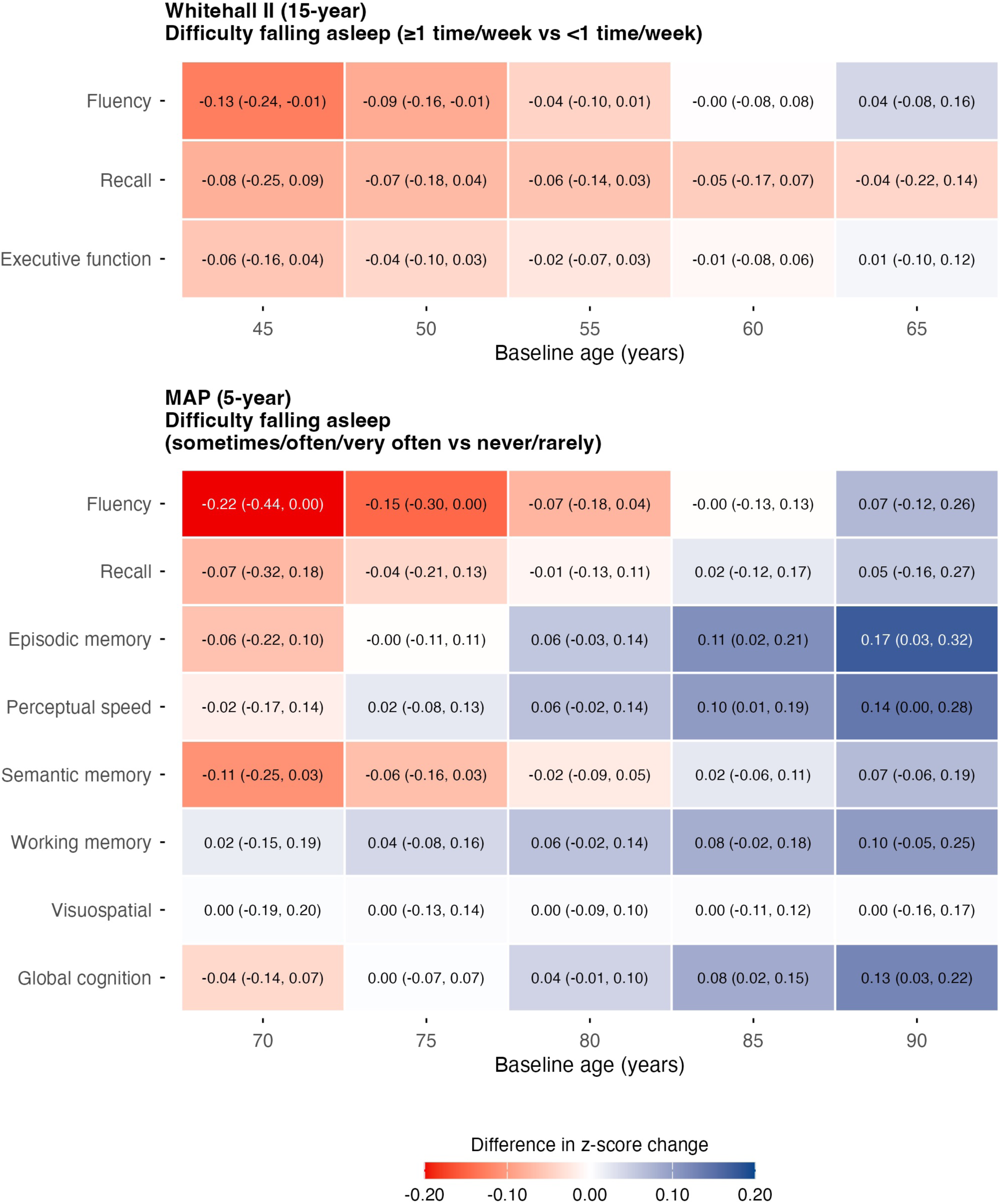
Differences in cognitive decline associated with difficulty falling asleep by baseline age in Whitehall II and MAP, Model 3. Heatmaps show β (95% CI) for differences in cognitive change associated with difficulty falling asleep (≥1 time/week vs <1 time/week for Whitehall II, sometimes/often/very often vs never/rarely for MAP). Estimates represent 15-year cognitive change in Whitehall II (baseline ages 45–65 years) and 5-year change in MAP (baseline ages 70–90 years). Estimates are derived from linear mixed-effects models including three-way interactions between baseline age, sleep, and follow-up time, with all lower-order terms, adjusted for sex, race and ethnicity, education, marital status, employment status (only for Whitehall II), body mass index, hypertension, diabetes, cardiovascular disease, depressive symptoms, smoking, alcohol use, physical activity, sleep medication and APOE ε4 carrier status. Random intercepts and slopes were included. Colours indicate the magnitude and direction of associations. Positive values (blue) indicate relatively less cognitive decline compared with the reference sleep category; negative values (red) indicate relatively greater decline.

In MAP, which primarily comprised older adults, five-year fluency differences became more positive with increasing baseline age, from −0·22 SD (95% CI −0·44 to 0·00) at age 70 to 0·07 SD (−0·12 to 0·26) at age 90 in Model 3 (**Figure 3**). The linear interaction for fluency and the quadratic interaction for episodic memory was significant across Models 1–3 (**Table S9**). Similar gradients were observed for several other domains, including perceptual speed and semantic memory, although their individual three-way interaction terms were not significant in Model 3.

### Sensitivity analysis

Age-dependent associations persisted after excluding 420 ELSA participants with incident dementia and using alternative categorisations of insomnia symptoms (<1, 1–2, ≥3 times/week) and sleep duration (≤6·4, 6·5–8·4, ≥8·5 hours) (**Table S8**). For difficulty falling asleep, the age-dependent gradient was more pronounced for 1–2 times/week, with larger effect sizes than the main binary analysis, whereas the ≥3 times/week group showed a similar but attenuated pattern (**Figures S6, S7**), suggesting that the age-dependent signal may not be dose-dependent and may be diluted by heterogeneous causes. For short sleep, the age-dependent pattern with fluency was consistent with findings in the main analysis, significant in Model 1 but attenuated after further adjustment.

Bayesian joint models yielded directionally consistent interaction estimates across all three cohorts (**Table S10**). The linear interactions for fluency and recall and the quadratic interaction for recall in ELSA, together with the quadratic interaction for fluency in Whitehall II, remained supported, with 95% credible intervals excluding zero. In MAP, the linear fluency interaction remained positive but was attenuated.

## Discussion

In this analysis of more than 16,000 adults across three large longitudinal cohorts spanning midlife to the oldest old, associations between sleep duration and insomnia symptoms and cognitive trajectories varied systematically by baseline age. Difficulty falling asleep showed robust age-dependent associations with cognitive trajectories: faster decline in midlife, attenuation through later adulthood, and reversal in advanced age. These associations persisted after adjustment for health-related and lifestyle covariates and were replicated across cohorts with complementary age coverage. Short sleep duration showed a similar gradient that attenuated after further covariate adjustment. Long sleep duration and nocturnal awakenings showed no age modification. Together, these findings suggest that the implications of sleep disturbances for cognitive decline may differ across stages of life.

These results help explain previously inconsistent evidence on sleep and cognitive ageing. In midlife, short sleep^10–12^ and difficulty falling asleep^11^ have been more consistently associated with worse cognitive outcomes and increased dementia risk. In population-based cohorts of adults aged ≥65 years (mean or median age > 70 years), these associations weaken or invert: insomnia was unrelated to cognitive decline according to a study in the MRC CFAS (UK)^15^ and Udine (Italy) cohorts ^16^, whereas difficulty maintaining sleep was paradoxically associated with reduced dementia risk in the Three-City (France)^14^ and NHATS (US) cohorts.^17^ Among women aged 80 and older, more insomnia complaints were associated with preserved cognitive function.^18^ Conversely, increased sleep propensity has been associated with adverse cognitive outcomes in these populations: excessive daytime sleepiness was associated with cognitive decline^15,16^ and with longitudinal amyloid accumulation^23^ in non-demented older adults, and in the oldest old, an increasing 24-hour sleepiness profile was associated with increased dementia risk.^24^ The few studies formally testing age as a modifier were limited by cross-sectional designs,^21^ binary age stratification,^19,20^ or restriction to individuals with subjective cognitive decline,^20^ all with modest sample sizes and incompletely covered age ranges. Our results extend these observations by showing that sleep–cognition associations shift continuously across baseline age, from adverse in midlife to attenuated or reversed in later life, in a pattern consistent across three independent cohorts.

In midlife, when overt neurodegenerative pathology remains relatively uncommon, sleep disturbances might primarily reflect upstream risk processes. Prolonged wakefulness and sleep fragmentation might lead to sustained neuronal activation and downstream neurodegenerative processes, while disrupted sleep might compromise cellular maintenance, metabolic homeostasis, and repair mechanisms.^3^ The attenuation of the short sleep association after adjustment for lifestyle and comorbid factors suggests that this relationship might be partly explained by behavioural and health-related factors. By contrast, the association with difficulty falling asleep persisted after adjustment, consistent with mechanisms not fully captured by conventional lifestyle and health factors, potentially reflecting central hyperarousal^25^ and its downstream neurobiological effects.

In later life, sleep–wake regulation depends on a balance between wake-promoting and sleep-promoting neural systems^26^ that becomes progressively disrupted with age and neurodegeneration. Normal ageing is associated with heightened excitability of arousal-promoting circuits,^27^ whereas advancing neurodegeneration, particularly of the locus coeruleus–noradrenergic system, might reduce arousability and increase sleep propensity.^9^ In this context, fewer insomnia complaints might no longer indicate preserved sleep health but rather diminished capacity of sleep-wake regulation circuits to generate and sustain wakefulness. Our finding that difficulty falling asleep at advanced ages was associated with less rather than more cognitive decline is consistent with this framework.

The age-dependent pattern was not uniform across sleep measures. Only difficulty falling asleep, which more directly reflects reduced sleep propensity at sleep onset, showed significant age modification. Nocturnal awakenings, by contrast, are frequently secondary to comorbid medical conditions such as sleep-disordered breathing, pain, and nocturia. These factors might fragment sleep and impair cognition from midlife^28^ through later life,^24^ potentially explaining why nocturnal awakenings showed no age-dependent gradient. Long sleep showed no age modification, likely because self-reported long sleep reflects a heterogeneous mix of behavioural, psychosocial, and medical causes, limiting its ability to capture the age-dependent signal that objective measures of sleep propensity have detected.^24^

These findings have potential implications across clinical and public health settings. In midlife, difficulty falling asleep was associated with approximately 0·12 SD less favourable 5-year change in word recall, an effect size comparable to recent multidomain prevention trials.^29^ Midlife sleep-onset difficulty might therefore represent a target for early intervention. Future trials could evaluate whether targeting insomnia in midlife can slow cognitive decline or delay dementia onset. In advanced age, the absence or resolution of insomnia complaints may not signal improved sleep health but rather a shift along the sleepiness–alertness continuum toward reduced arousal. Comprehensive assessment encompassing both insomnia symptoms and markers of increased sleep propensity, alongside routine cognitive monitoring, may be needed to fully characterise sleep-related cognitive risk in older adults.

This study has several strengths: a large, longitudinal design across three independent cohorts in the UK and US; broad and complementary age ranges from midlife to late life, including a nationally representative sample; harmonised sleep and cognition measures enabling direct cross-cohort comparison of age-dependent associations; and a modelling framework that formally tests effect modification by age.

Several limitations warrant consideration. First, sleep measures were self-reported and subject to misclassification, particularly among individuals with cognitive impairment.^30^ However, age-dependent patterns persisted after excluding participants who developed dementia during follow-up. Subjective sleep nonetheless has unique value in reflecting symptom burden and healthcare-seeking behaviour. Future studies integrating objective measures are needed. Second, despite harmonisation efforts, residual measurement heterogeneity cannot be excluded. The absence of biomarker data further limits inference regarding underlying neuropathology. Third, age at sleep report may partly reflect birth cohort, though consistent patterns across ELSA and WHII—whose participants share overlapping birth cohorts (median birth year 1943) but were assessed at different ages—argue against this explanation. Fourth, selective survival and differential attrition may partly contribute to the apparent reversal at older ages. Joint models yielded directionally consistent effect estimates, providing some reassurance regarding informative attrition. Even if selection contributes to the age gradient, the findings highlight the limits of extrapolating midlife sleep–cognition associations to older populations. Finally, this study was not designed to estimate causal effects, and the predominantly White study population may limit generalisability.

In conclusion, the cognitive implications of sleep disturbances shift across the adult lifespan. Difficulty falling asleep and short sleep are associated with faster cognitive decline in midlife; in later life, these characteristics are instead associated with relatively less decline. These results identify difficulty falling asleep in midlife as a candidate modifiable target for dementia prevention and reframe the interpretation of sleep complaints in advanced age as potential markers of declining arousal rather than benign ageing. Midlife may represent a critical window in which sleep-targeted interventions could be evaluated to preserve long-term cognitive health.

## Contributors

YF and YL conceived and designed the study. YF, AQ, and NO conducted the statistical analyses and directly accessed and verified the underlying data reported in the manuscript. YF drafted the manuscript. YL acquired funding and supervised the study. All authors contributed to interpretation of the results, critically revised the manuscript for important intellectual content, and approved the final version. All authors had final responsibility for the decision to submit for publication.

## Declaration of interests

We declare no competing interests.

## Data Sharing

Data from the ELSA and WHII are available through the UK Dementias Platform and can be accessed upon reasonable request and with appropriate approvals. Data from the Rush MAP are available through the Rush Alzheimer’s Disease Centre Resource Sharing Hub at www.radc.rush, subject to data use agreements and institutional approvals.

## Data Availability

Data from the ELSA and WHII are available through the www.dementiasplatform.uk and can be accessed upon reasonable request and with appropriate approvals. Data from the Rush MAP are available through the Rush Alzheimer's Disease Centre Resource Sharing Hub at www.radc.rush, subject to data use agreements and institutional approvals.

## Acknowledgment

This work was supported by the US National Institute on Aging (R01AG083836). We thank the participants and study teams of the English Longitudinal Study of Ageing, Whitehall II study, and the Rush Memory and Aging Project for their contributions. The Rush Memory and Aging Project is supported by the US National Institute on Aging (R01AG17917).

## Notes

### Competing Interest Statement

The authors have declared no competing interest.

### Author Declarations

Data from the ELSA and WHII are available through the www.dementiasplatform.uk and can be accessed upon reasonable request and with appropriate approvals. Data from the Rush MAP are available through the Rush Alzheimer's Disease Centre Resource Sharing Hub at www.radc.rush, subject to data use agreements and institutional approvals. All datasets used in this study were de-identified prior to their use in the study.

## References

1 Mander BA, Winer JR, Walker MP. Sleep and Human Aging. Neuron 2017; 94: 19–36.

2 Dohm-Hansen S, English JA, Lavelle A, Fitzsimons CP, Lucassen PJ, Nolan YM. The ‘middle-aging’ brain. Trends in Neurosciences 2024; 47: 259–72.

3 Parhizkar S, Holtzman DM. The night’s watch: Exploring how sleep protects against neurodegeneration. Neuron 2025; 113: 817–37.

4 Dubois B, Hampel H, Feldman HH, et al. Preclinical Alzheimer’s disease: Definition, natural history, and diagnostic criteria. Alzheimers Dement 2016; 12: 292–323.

5 Van Egroo M, Koshmanova E, Vandewalle G, Jacobs HIL. Importance of the locus coeruleus-norepinephrine system in sleep-wake regulation: Implications for aging and Alzheimer’s disease. Sleep Medicine Reviews 2022; 62: 101592.

6 Overton M, Sindi S, Basna R, Elmståhl S. Excessive sleep is associated with worse cognition, cognitive decline, and dementia in mild cognitive impairment. *Alzheimer’s & Dementia: Diagnosis*, Assessment & Disease Monitoring 2025; 17: e70093.

7 Livingston G, Huntley J, Liu KY, et al. Dementia prevention, intervention, and care: 2024 report of the Lancet standing Commission. The Lancet 2024; **404**: 572–628.

8 Aarsland D, Sunde AL, Tovar-Rios DA, et al. Prevalence of Alzheimer’s disease pathology in the community. Nature 2025;: 1–5.

9 Oh JY, Walsh CM, Ranasinghe K, et al. Subcortical Neuronal Correlates of Sleep in Neurodegenerative Diseases. JAMA Neurol 2022; 79: 498–508.

10 Sabia S, Fayosse A, Dumurgier J, et al. Association of sleep duration in middle and old age with incidence of dementia. Nat Commun 2021; 12: 2289.

11 Tan X, Åkerstedt T, Lagerros YT, et al. Interactive association between insomnia symptoms and sleep duration for the risk of dementia—a prospective study in the Swedish National March Cohort. Age and Ageing 2023; 52: afad163.

12 Ell J, Schiel JE, Feige B, et al. Sleep health dimensions and shift work as longitudinal predictors of cognitive performance in the UK Biobank cohort. SLEEP 2023; 46: zsad093.

13 Scullin MK, Bliwise DL. Sleep, Cognition, and Normal Aging: Integrating a Half Century of Multidisciplinary Research. Perspect Psychol Sci 2015; 10: 97–137.

14 Jaussent I, Bouyer J, Ancelin M-L, et al. Excessive Sleepiness is Predictive of Cognitive Decline in the Elderly. Sleep 2012; 35: 1201–7.

15 Keage HAD, Banks S, Yang KL, Morgan K, Brayne C, Matthews FE. What sleep characteristics predict cognitive decline in the elderly? Sleep Medicine 2012; 13: 886–92.

16 Merlino G, Piani A, Gigli GL, et al. Daytime sleepiness is associated with dementia and cognitive decline in older Italian adults: A population-based study. Sleep Medicine 2010; 11: 372–7.

17 Wong R, Lovier MA. Sleep Disturbances and Dementia Risk in Older Adults: Findings From 10 Years of National U.S. Prospective Data. Am J Prev Med 2023; 64: 781–7.

18 Goveas JS, Rapp SR, Hogan PE, et al. Predictors of Optimal Cognitive Aging in 80+ Women: The Women’s Health Initiative Memory Study. J Gerontol A Biol Sci Med Sci 2016; 71 **Suppl 1**: S62–71.

19 Arévalo SP, Nguyen-Rodriguez ST, Scott TM, Gao X, Falcón LM, Tucker KL. Longitudinal Associations Between Sleep and Cognitive Function in a Cohort of Older Puerto Rican Adults: Sex and Age Interactions. J Gerontol A Biol Sci Med Sci 2023; 78: 1816–25.

20 Huang J, Perrin NA, Spira AP, et al. Sleep disturbance and cognitive trajectories among older adults with subjective cognitive decline: the roles of age and sleep treatment. Sleep 2025;: zsaf234.

21 Cohen DE, Kim H, Levine A, Devanand DP, Lee S, Goldberg TE. Effects of age on the relationship between sleep quality and cognitive performance: Findings from the Human Connectome Project-Aging cohort. International Psychogeriatrics 2024; 36: 1171–81.

22 Brookes ST, Whitely E, Egger M, Smith GD, Mulheran PA, Peters TJ. Subgroup analyses in randomized trials: risks of subgroup-specific analyses;: power and sample size for the interaction test. Journal of Clinical Epidemiology 2004; 57: 229–36.

23 Tu Y-Y, Cui L, Zhang Z, et al. Subjective sleep characteristics in non-demented adults: domain-specific associations with multimodal Alzheimer’s disease biomarkers. Alzheimer’s & Dementia 2025; 21: e70932.

24 Milton S, Cavaillès C, Ancoli-Israel S, Stone KL, Yaffe K, Leng Y. Five-Year Changes in 24-Hour Sleep-Wake Activity and Dementia Risk in Oldest Old Women. Neurology 2025; 104: e213403.

25 Fernandez-Mendoza J. Insomnia Phenotypes, Cardiovascular Risk and Their Link to Brain Health. Circulation Research 2025; 137: 727–45.

26 Saper CB, Fuller PM, Pedersen NP, Lu J, Scammell TE. Sleep State Switching. Neuron 2010; 68: 1023–42.

27 Li S-B, Damonte VM, Chen C, et al. Hyperexcitable arousal circuits drive sleep instability during aging. Science 2022; 375: eabh3021.

28 Leng Y, Knutson K, Carnethon MR, Yaffe K. Association Between Sleep Quantity and Quality in Early Adulthood With Cognitive Function in Midlife. Neurology 2024; 102: e208056.

29 Baker LD, Espeland MA, Whitmer RA, et al. Structured vs Self-Guided Multidomain Lifestyle Interventions for Global Cognitive Function: The US POINTER Randomized Clinical Trial. JAMA 2025; 334: 681–91.

30 Most EIS, Aboudan S, Scheltens P, Van Someren EJW. Discrepancy Between Subjective and Objective Sleep Disturbances in Early- and Moderate-Stage Alzheimer Disease. The American Journal of Geriatric Psychiatry 2012; 20: 460–7.

